# Circulating Fatty Acid Synthase and Modified Frailty Index-5 Are Additive Predictors of Adverse Outcomes After Elective Vascular Surgery

**DOI:** 10.64898/2026.08.10.26360144

**Authors:** Mohamed Zaghloul, Ryan Catlett, Bera Koklu, Abdullah Elahi, Omar Soltan, Jad Yacoub, Dina Ibrahim, Wahid Abu-Amer, Feng Gao, Mohamed A Zayed

## Abstract

**Background:** Preoperative risk assessment in vascular surgery relies on clinical scores and lipids that do not capture atherosclerotic disease activity. Circulating fatty acid synthase (cFAS) is a liver-derived enzyme whose concentration correlates with arterial plaque FAS content independent of LDL. The 5-item modified frailty index (mFI-5) is a validated predictor of postoperative mortality. Whether cFAS predicts outcomes after vascular surgery, and whether combining it with the mFI-5 improves risk discrimination, have not been examined.

**Methods:** We studied 657 patients undergoing elective vascular surgery at a single center (2014 to 2023). cFAS was classified as non-detectable (n = 306) or, among detectable values, by tertiles (n = 117 each). Multivariable Cox models assessed associations with major adverse events (MAE), major adverse cardiovascular events (MACE), major adverse limb events (MALE), reintervention, and mortality, and Harrell’s C-statistic quantified the incremental discrimination gained by adding cFAS and the mFI-5 to standard clinical covariates.

**Results:** High serum cFAS was independently associated with 5-year MAE (adjusted hazard ratio [aHR] 1.94; 95% CI 1.31-2.85), mortality (aHR 1.77; 1.05 to 3.00), MALE (aHR 4.53; 2.04 to 10.05), and reintervention (aHR 2.50; 1.37 to 4.57), but not MACE. Severe frailty (mFI-5 of 3 or higher) was associated with MACE (aHR 2.69; 1.29 to 5.58) and MAE (aHR 2.46; 1.30 to 4.65) but not limb endpoints at 1 year. Adding cFAS raised the 1-year MALE C-statistic from 0.649 to 0.764; the combined model yielded the highest discrimination.

**Conclusions:** cFAS and mFI-5 were independently and additively associated with adverse outcomes after elective vascular surgery. cFAS was associated with limb events and mortality, the mFI-5 with cardiovascular events. Combining them improved discrimination over standard covariates.

**CLINICAL PERSPECTIVE:** *What Is New?:*

- A single preoperative measurement of circulating fatty acid synthase (cFAS) independently predicted adverse outcomes after elective vascular surgery, establishing cFAS as a prognostic biomarker of atherosclerotic disease activity beyond its previously reported diagnostic role.
- cFAS and the 5-item modified frailty index (mFI-5) captured complementary and additive dimensions of postoperative risk: disease activity and physiologic reserve, respectively.
- Combining cFAS with the mFI-5 improved risk discrimination beyond standard clinical covariates for all endpoints examined.

*What Are the Clinical Implications?:*

- cFAS may identify residual atherosclerotic risk not captured by standard lipid panels, offering a novel dimension of preoperative assessment in vascular surgery.
- Incorporating a routine frailty assessment (mFI-5) alongside a disease-activity biomarker (cFAS) could enhance preoperative risk stratification by addressing two distinct and additive determinants of surgical outcome.
- Prospective external validation is warranted before cFAS is adopted as an adjunct to established preoperative cardiovascular risk assessment in vascular surgery.

## INTRODUCTION

Atherosclerotic cardiovascular disease (ASCVD) is a leading cause of morbidity and mortality worldwide and underlies the majority of the estimated $393 billion in annual cardiovascular health care costs in the United States, costs projected to nearly quadruple by 2050.^1,2^ In patients undergoing vascular surgery, preoperative risk stratification relies on clinical risk scores, such as the 10-year ASCVD risk estimate, and on standard lipids, mainly low-density lipoprotein (LDL).^3^ These measures estimate risk at the population level, but they reflect the burden of risk factors rather than the activity of the disease itself.^3^ In a collaborative analysis of more than 31,000 statin-treated patients, the residual risk of cardiovascular events and death was driven by ongoing inflammation rather than by LDL.^4^ Once LDL is controlled, the risk that remains therefore reflects the activity of the disease rather than the lipid burden.^4^ The CANTOS trial established that this risk is modifiable. Anti-inflammatory therapy with canakinumab reduced recurrent cardiovascular events by 15% without lowering LDL, evidence that atherosclerotic disease activity drives these events independently of the lipid burden.^5^ Together, these findings point to the need for a circulating biomarker that reflects disease activity directly.

Circulating fatty acid synthase (cFAS) is a liver-derived serum enzyme, bound to LDL particles, that was first identified in patients with diabetes and carotid artery stenosis.^6^ FAS catalyzes the *de novo* synthesis of saturated fatty acids, and in macrophages its activity organizes the plasma membrane for inflammatory signaling.^7^ In peripheral atherosclerosis, cFAS rises with disease severity and correlates with the FAS content of arterial plaque, independent of diabetes and smoking.^8^ These associations suggest that cFAS may contribute to atheroprogression rather than passively reflecting disease severity. Consistent with this, pharmacologic inhibition of FAS in mice reduces macrophage foam cell formation and aortic atherosclerosis without altering circulating cholesterol.^9^ cFAS was recently shown to distinguish patients with peripheral arterial disease (PAD) from healthy controls, and to further discriminate PAD from chronic limb-threatening ischemia (CLTI), and was validated as an independent diagnostic biomarker.^10^ In each of these studies, however, cFAS was assessed cross-sectionally as a diagnostic test, and whether a preoperative measurement predicts outcomes after vascular surgery has not been examined.^6,8,10^

In addition to disease activity, physiologic reserve is a distinct determinant of surgical outcome. This reserve is captured by the 5-item modified frailty index (mFI-5), a measure derived from the American College of Surgeons National Surgical Quality Improvement Program (ACS NSQIP) that is calculated from five routinely documented comorbidities and predicts postoperative mortality and complications across surgical specialties.^11^ In a meta-analysis of 111 studies and more than 25 million surgical patients, the mFI-5 outperformed the older Charlson Comorbidity Index in predicting short-term postoperative mortality and major complications.^12^ The mFI-5 requires fewer variables than the original 11-item index yet shows comparable or better predictive performance for adverse outcomes.^13^ In vascular surgery, frailty ranks among the strongest predictors of long-term survival; in a Vascular Quality Initiative (VQI)-Medicare linked cohort of more than 109,000 patients undergoing elective vascular procedures, greater frailty was associated with a nearly sixfold higher 5-year mortality.^14^ Frailty, however, reflects systemic reserve and comorbidity rather than the activity of the underlying atherosclerotic disease.^11,14^

In other surgical settings, adding a serum biomarker to a clinical risk score has improved outcome prediction. Preoperative N-terminal pro-B-type natriuretic peptide (NT-proBNP) added to the Revised Cardiac Risk Index improved prediction of cardiovascular events after noncardiac surgery, and a panel of serum biomarkers added to an established clinical score improved prediction of complications after cardiac surgery.^15,16^ These biomarkers, however, report myocardial stress or systemic processes rather than the activity of the atherosclerotic plaque. cFAS and the mFI-5 may instead capture two distinct and additive components of postoperative risk: the activity of the disease and the patient’s physiologic reserve. We therefore hypothesized that preoperative cFAS and mFI-5 independently and additively predict short-term and long-term adverse outcomes after vascular surgery, and that combining them improves risk discrimination beyond standard clinical covariates.

## METHODS

### Study Design and Population

This single-center retrospective cohort study examined consecutive patients who underwent elective vascular surgery at Washington University in St. Louis between September 2014 and February 2023. All patients were prospectively enrolled in an institutional vascular surgery biobank at the time of their index procedure, with preoperative serum collected for biomarker quantification. All patients with a valid fasting serum cFAS measurement (detectable or non-detectable) were included in the analysis (n = 657).

Patients were classified by primary surgical indication into 4 mutually exclusive groups: PAD/CLTI, carotid artery disease, aortic disease, and other vascular conditions (**Table 1**). Classification was determined by the attending vascular surgeon at biobank enrollment and verified by chart review. Clinical outcomes were ascertained through systematic electronic health record review, with follow-up extending through May 2026.

**Table 1.** Baseline Characteristics Stratified by cFAS Detectability and cFAS Tertile.

|  | Total<br>(N=657) | Non-Detectable<br>cFAS<br>(n = 306) | Detectable cFAS, pg/mg (n = 351) |  |  | P<br>(4-grp) | P<br>(det) |
| --- | --- | --- | --- | --- | --- | --- | --- |
|  |  |  | Low<br>(≤265, n = 117) | Mid<br>(266–748, n = 117) | High<br>(>748, n = 117) |  |  |
| Demographics |  |  |  |  |  |  |  |
| Age, years | 63.2±14.4 | 62.2±16.0 | 64.5±12.2 | 64.1±14.5 | 63.8±11.7 | 0.379 | 0.087 |
| Male | 398 (60.6) | 174 (56.9) | 70 (59.8) | 77 (65.8) | 77 (65.8) | 0.214 | 0.069 |
| White | 559 (85.3) | 264 (86.8) | 104 (88.9) | 99 (84.6) | 92 (78.6) | 0.112 | 0.313 |
| BMI, kg/m² | 28.1±6.5 | 27.7±6.4 | 28.5±6.0 | 28.5±6.8 | 28.5±6.9 | 0.530 | 0.137 |
| Current smoker | 245 (37.5) | 100 (32.9) | 46 (39.7) | 45 (38.5) | 54 (46.2) | 0.080 | 0.025 |
| Comorbidities |  |  |  |  |  |  |  |
| Hypertension | 520 (79.1) | 239 (78.1) | 91 (77.8) | 94 (80.3) | 96 (82.1) | 0.793 | 0.539 |
| Diabetes mellitus | 219 (33.3) | 61 (19.9) | 35 (29.9) | 61 (52.1) | 62 (53.0) | <0.001 | <0.001 |
| Hyperlipidemia | 442 (73.4) | 196 (72.3) | 85 (77.3) | 79 (70.5) | 82 (75.2) | 0.645 | 0.581 |
| CAD | 254 (43.8) | 105 (41.8) | 46 (42.2) | 46 (40.4) | 57 (53.8) | 0.148 | 0.406 |
| MI | 117 (21.6) | 50 (21.2) | 23 (22.1) | 23 (21.9) | 21 (21.6) | 0.997 | 0.842 |
| CHF | 101 (15.4) | 41 (13.4) | 13 (11.1) | 18 (15.4) | 29 (24.8) | 0.015 | 0.190 |
| COPD | 166 (25.3) | 74 (24.2) | 26 (22.2) | 30 (25.6) | 36 (30.8) | 0.449 | 0.551 |
| CKD | 145 (31.7) | 75 (37.5) | 24 (27.3) | 17 (20.0) | 29 (34.1) | 0.023 | 0.018 |
| Stroke | 101 (21.6) | 45 (22.0) | 20 (22.7) | 19 (20.0) | 17 (21.3) | 0.973 | 0.864 |
| Dementia | 10 (2.2) | 6 (3.1) | 3 (3.4) | 0 (0.0) | 1 (1.3) | 0.297 | 0.340 |
| Clinical Presentation |  |  |  |  |  |  |  |
| PAD/CLTI | 221 (33.7) | 66 (21.6) | 35 (30.2) | 55 (47.4) | 65 (55.6) | <0.001 | <0.001 |
| Carotid stenosis | 204 (31.1) | 105 (34.3) | 40 (34.5) | 33 (28.4) | 26 (22.2) |  |  |
| Aortic disease | 171 (26.1) | 100 (32.7) | 30 (25.9) | 18 (15.5) | 23 (19.7) |  |  |
| Other vascular | 59 (9.0) | 35 (11.4) | 11 (9.5) | 10 (8.6) | 3 (2.6) |  |  |
| **Surgical History** |  |  |  |  |  |  |  |
| Leg revascularization | 129 (19.6) | 37 (12.1) | 21 (17.9) | 34 (29.1) | 37 (31.6) | <0.001 | <0.001 |
| Amputation | 27 (4.1) | 6 (2.0) | 2 (1.7) | 10 (8.5) | 9 (7.7) | 0.002 | 0.010 |
| Carotid intervention | 69 (10.5) | 27 (8.8) | 19 (16.2) | 16 (13.7) | 7 (6.0) | 0.032 | 0.190 |
| Aneurysm repair | 39 (5.9) | 19 (6.2) | 7 (6.0) | 5 (4.3) | 8 (6.8) | 0.884 | 0.869 |
| CABG | 116 (17.7) | 50 (16.3) | 20 (17.1) | 26 (22.2) | 20 (17.1) | 0.553 | 0.409 |
| **Medications** |  |  |  |  |  |  |  |
| Aspirin | 441 (67.1) | 200 (65.4) | 81 (69.2) | 83 (70.9) | 77 (65.8) | 0.675 | 0.369 |
| Statin | 443 (67.4) | 202 (66.0) | 80 (68.4) | 84 (71.8) | 77 (65.8) | 0.687 | 0.470 |
| Beta blocker | 308 (46.9) | 139 (45.4) | 52 (44.4) | 58 (49.6) | 59 (50.4) | 0.689 | 0.485 |
| ACE inhibitor/ARB | 280 (42.6) | 130 (42.5) | 43 (36.8) | 56 (47.9) | 51 (43.6) | 0.390 | 0.948 |
| Anticoagulant | 111 (16.9) | 46 (15.0) | 22 (18.8) | 20 (17.1) | 23 (19.7) | 0.637 | 0.234 |
| Insulin | 97 (14.8) | 28 (9.2) | 19 (16.2) | 20 (17.1) | 30 (25.6) | <0.001 | <0.001 |
| Metformin | 95 (14.5) | 28 (9.2) | 17 (14.5) | 30 (25.6) | 20 (17.1) | <0.001 | <0.001 |
| **Laboratory Values** |  |  |  |  |  |  |  |
| Creatinine, mg/dL | 1.0±0.3 | 1.0±0.3 | 1.0±0.3 | 1.0±0.3 | 1.0±0.3 | 0.942 | 0.903 |
| eGFR, mL/min/1.73m<sup>2</sup> | 78.0±21.7 | 78.0±20.9 | 77.0±22.4 | 78.8±21.2 | 78.3±23.7 | 0.946 | 0.991 |
| HbA1c, % | 7.0±1.6 | 6.6±1.3 | 7.2±2.3 | 6.9±1.3 | 7.5±1.7 | 0.043 | 0.017 |
| Total cholesterol, mg/dL | 143.2±42.1 | 147.1±41.9 | 143.9±37.5 | 140.8±45.0 | 140.3±45.9 | 0.784 | 0.384 |
| LDL, mg/dL | 80.8±33.0 | 82.9±32.0 | 82.7±30.3 | 78.0±31.3 | 78.6±39.6 | 0.738 | 0.544 |
| HDL, mg/dL | 39.8±12.9 | 43.5±14.2 | 39.8±10.2 | 39.1±12.7 | 36.2±14.1 | 0.015 | 0.006 |
| Triglycerides, mg/dL | 141.3±97.8 | 121.7±70.8 | 135.8±69.1 | 152.7±140.7 | 159.5±99.9 | 0.118 | 0.058 |
| **Frailty** |  |  |  |  |  |  |  |
| mFI-5 | 1.7±1.1 | 1.5±1.0 | 1.6±1.0 | 1.9±1.2 | 2.1±1.2 | <0.001 | <0.001 |
| Non-frail (0) | 97 (14.8) | 51 (16.7) | 18 (15.4) | 17 (14.5) | 11 (9.4) | 0.001 | 0.001 |
| Pre-frail (1) | 181 (27.5) | 101 (33.0) | 35 (29.9) | 19 (16.2) | 26 (22.2) |  |  |
| Frail (2) | 234 (35.6) | 103 (33.7) | 41 (35.0) | 50 (42.7) | 40 (34.2) |  |  |
| Severely frail ($\geq 3$) | 145 (22.1) | 51 (16.7) | 23 (19.7) | 31 (26.5) | 40 (34.2) | | |

Data are presented as mean ± SD or n (%). Percentages are calculated among patients with available data for each variable. P (4- grp), comparison across all four groups (ANOVA for continuous, chi-squared or Fisher's exact for categorical); P (det), non- detectable vs detectable (t-test for continuous, chi-squared or Fisher's exact for categorical). BMI, body mass index; CABG, coronary artery bypass grafting; CAD, coronary artery disease; CHF, congestive heart failure; CKD, chronic kidney disease; COPD, chronic obstructive pulmonary disease; eGFR, estimated glomerular filtration rate; HbA1c, hemoglobin A1c; HDL, high- density lipoprotein; LDL, low-density lipoprotein; mFI-5, modified frailty index-5; MI, myocardial infarction; cFAS, circulating fatty acid synthase.

The study was approved by the Washington University in St. Louis School of Medicine Institutional Review Board. All patients provided written informed consent for inclusion in the prospectively maintained institutional vascular surgery registry and serum biobank.

### Blood Collection and cFAS Quantification

Serum collection and processing were performed as previously described.^10^ Intravenous whole blood samples were obtained from fasting patients (minimum of 6 hours) prior to elective vascular surgery (**Table 1**). Samples were collected in red- and green-topped vacutainer tubes and centrifuged to isolate serum and plasma components. Serum was aliquoted into 100 μL fractions and stored at −80°C for subsequent analysis.

cFAS concentration was measured using a commercially available enzyme-linked immunosorbent assay (ELISA; Aviva Systems Biology, catalog no. OKEH01027) with a quantification range of 312 to 20,000 pg/mL. To account for inter-patient variation in the duration of preoperative fasting, total serum protein concentration was determined by Bradford assay. Raw cFAS values were normalized to total protein content as cFAS (pg/mg) = cFAS (pg/mL) ÷ total protein (mg/mL), yielding normalized cFAS values in pg/mg.

### cFAS Categorization

Of 657 enrolled patients, 351 (53.4%) had cFAS concentrations within the quantification range of the ELISA assay and were classified as detectable. The remaining 306 patients (46.6%) had cFAS levels below the lower limit of detection and were classified as non-detectable.

Within the 351 patients with detectable cFAS, cFAS values were stratified into sample-based tertiles: Low (≤265 pg/mg, n = 117), Mid (266 to 748 pg/mg, n = 117), and High (>748 pg/mg, n = 117). The resulting four-group ordinal variable (Non-detectable, Low, Mid, High) served as the primary predictor specification. Sample-based quantile tertiles were chosen because no established clinical cutoffs exist for cFAS; applying value-based tertile boundaries to the right-skewed distribution among detectable samples (skewness 1.93; range 11.2 to 3,844 pg/mg) would have produced severely unequal group sizes.

### Modified Frailty Index-5

mFI-5 is a validated 5-item composite frailty measure scored from 0 to 5, originally derived from ACS NSQIP database.^11^ Its components are diabetes mellitus, hypertension, congestive heart failure, chronic obstructive pulmonary disease, and functional status (partially or totally dependent). Because direct functional status assessments were not recorded in the biobank registry, functional impairment was assessed using neurological and cognitive proxy variables: dementia, history of stroke, residual stroke deficits, and hemiplegia. Dementia, stroke, and hemiplegia are established contributors to functional dependence and are incorporated as prognostically weighted comorbidities in the Charlson Comorbidity Index.^17^ Patients were categorized as non-frail (score 0), pre-frail (1), frail (2), or severely frail (≥3), consistent with previously reported thresholds.^18^ For the combined cFAS–frailty analysis, patients were additionally dichotomized as low frailty (mFI-5 <2) or high frailty (mFI-5 ≥2), a threshold commonly used to define frailty in vascular surgery cohorts.^19^

### Outcome Definitions

Five time-to-event endpoints were analyzed. The primary endpoints were: (1) all-cause mortality; (2) major adverse cardiovascular events (MACE), defined as the composite of death, myocardial infarction, and stroke; (3) major adverse limb events (MALE), defined as above-ankle amputation of the index limb or acute limb ischemia requiring urgent revascularization; and (4) major adverse events (MAE), defined as the composite of MACE and MALE. The secondary endpoint was target-related reintervention, defined as any unplanned surgical or endovascular procedure performed on the same vascular territory as the index operation.

Time-to-event intervals were calculated from the date of the index procedure (time zero) to the date of the first qualifying event or last known follow-up, whichever occurred first. Patients who did not experience an event were censored at the date of last documented clinical contact.

### Statistical Analysis

Continuous variables were reported as mean ± standard deviation and were compared across the four cFAS groups using one-way analysis of variance. Categorical variables are presented as counts with percentages and were compared using the chi-squared test; Fisher’s exact test was substituted when any expected cell count was fewer than 5. A two-group comparison (detectable vs. non-detectable cFAS) was performed in parallel using the independent-samples t-test and chi-squared test, respectively.

To evaluate whether cFAS and mFI-5 capture distinct biological constructs, the correlation between the two predictors was assessed using the Spearman rank coefficient (cFAS vs. mFI-5 score) among patients with both measurements available.

Cox proportional hazards models were constructed for each endpoint at short-term (1-year) and long-term (5-year) time horizons. Unadjusted and covariate-adjusted hazard ratios (HR) with 95% confidence intervals (CI) are reported. The primary predictors were the four-group cFAS variable (reference: non-detectable) and mFI-5 category (reference: non-frail, score 0). Kaplan-Meier estimates with log-rank testing were used to compare event-free survival across cFAS groups and mFI-5 categories for each endpoint. Kaplan-Meier estimates were also generated for a four-group combined cFAS detectability–frailty classification, defined by cFAS detectability and binary frailty status (low frailty, mFI-5 <2 [non-frail or pre-frail]; high frailty, mFI-5 ≥2 [frail or severely frail]). Cumulative event rates at 1 and 5 years were derived from these curves to display the incidence of each outcome over time.

Covariates were pre-specified on clinical grounds: age, sex, smoking status (ever vs. never smoker), estimated glomerular filtration rate (eGFR), and body mass index (BMI). These variables were selected as established prognostic factors for adverse outcomes after elective vascular surgery that do not overlap with the components of either primary predictor. Age and sex are standard demographic confounders in cardiovascular outcome models. Smoking is the predominant modifiable risk factor for peripheral arterial disease progression and cardiovascular mortality.^20^ eGFR is an independent predictor of both perioperative and long-term mortality in vascular surgical populations.^21^ BMI accounts for metabolic and nutritional risk relevant to surgical outcomes.^22^ Because the mFI-5 already incorporates diabetes mellitus, hypertension, congestive heart failure, chronic obstructive pulmonary disease, and functional impairment, these conditions were not included as separate covariates to avoid collinearity.

Discriminatory performance was assessed using Harrell’s C-statistic across a series of progressively adjusted models: covariates only, covariates plus cFAS, covariates plus mFI-5, and covariates plus both predictors. The change in C-statistic (ΔC) relative to the covariates-only model is reported for each incremental addition to quantify the independent and joint predictive contributions of cFAS and mFI-5.

To determine whether the joint predictive effect of cFAS and mFI-5 is additive or synergistic, likelihood ratio tests were performed comparing additive Cox models (cFAS four-group + mFI-5 category) with multiplicative models incorporating the cFAS × mFI-5 interaction term (**Supplemental Table 1**). Interaction terms with zero events in specific strata were omitted by the model, and the degrees of freedom vary accordingly across endpoints and time horizons.

Sensitivity analyses examined alternative predictor specifications to evaluate the robustness of the primary categorical findings (**Supplemental Table 2**). These included: cFAS modeled as a continuous variable (per 100 pg/mg increase), natural log-transformed cFAS [ln(cFAS), per 1-unit increase on the log scale], mFI-5 modeled as a continuous score (per 1-point increase). Continuous and log-transformed cFAS models were restricted to patients with detectable cFAS (n = 351).

The proportional hazards assumption was evaluated for all adjusted models using scaled Schoenfeld residuals. All analyses were performed using Stata version 19.5 (StataCorp, College Station, TX). Two-sided P values < 0.05 were considered statistically significant.

Dr. Zaghloul and Dr. Zayed have full access to all study data and take responsibility for the integrity of the data and its analysis.

## RESULTS

### Study Population and Baseline Characteristics

Of 657 patients, 351 (53.4%) had detectable cFAS and 306 (46.6%) had non-detectable levels. Among patients with detectable cFAS, concentrations were divided into Low, Mid, and High tertiles (n = 117 each). The mean age was 63.2 ± 14.4 years, 398 (60.6%) were male, and 559 (85.3%) were White. The primary surgical indications were PAD/CLTI (n = 221, 33.7%), carotid disease (n = 204, 31.1%), aortic disease (n = 171, 26.1%), and other vascular diseases (n = 59, 9.0%) (**Table 1**).

Cardiovascular risk factors and markers of disease severity increased across cFAS groups. Diabetes mellitus rose from 19.9% in the non-detectable group to 53.0% in the High tertile (P < 0.001), with parallel increases in insulin (non-detectable, 9.2% vs. High, 25.6%; P < 0.001) and metformin use (non-detectable, 9.2% vs. High, 17.1%; P < 0.001), higher HbA1c (non-detectable, 6.6% vs. High, 7.5%; P = 0.043), and lower HDL cholesterol (non-detectable, 43.5 vs. High, 36.2 mg/dL; P = 0.015). Current smoking was more frequent at higher cFAS levels (non-detectable, 32.9% vs. High, 46.2%), reaching significance in the detectable-versus-non-detectable comparison (P = 0.025) but not across the four groups (P = 0.080).

The proportion of patients whose primary surgical indication was PAD/CLTI rose across cFAS groups, from 21.6% (non-detectable) to 30.2% (Low), 47.4% (Mid), and 55.6% (High) (P < 0.001). Prior lower-extremity revascularization (non-detectable, 12.1% vs. High, 31.6%; P < 0.001) and prior amputation (non-detectable, 2.0% vs. High, 7.7%; P = 0.002) were also more common at higher cFAS levels.

There were no significant differences in age, sex, BMI, or the prevalence of hypertension, coronary artery disease, or stroke across groups (all P > 0.05). Medication use (aspirin, beta blockers, ACE inhibitors/ARBs, and anticoagulants) was similar across groups (all P > 0.05).

Hyperlipidemia prevalence and the lipid profile did not differ across cFAS groups. Hyperlipidemia was equally common (non-detectable, 72.3% vs. High, 75.2%; P = 0.65), statin use was similar (66.0% vs. 65.8%; P = 0.69), and LDL (82.9 vs. 78.6 mg/dL; P = 0.74), total cholesterol (147.1 vs. 140.3 mg/dL; P = 0.78), and triglycerides (121.7 vs. 159.5 mg/dL; P = 0.12) were comparable. cFAS therefore rose independently of LDL and the broader lipid profile (**Table 1**).

### Frailty Distribution

Among 657 patients with available mFI-5 data, the mean score was 1.7 ± 1.1. Overall, 14.8% were non-frail (score 0), 27.5% pre-frail (score 1), 35.6% frail (score 2), and 22.1% severely frail (score ≥ 3). The mean mFI-5 increased across cFAS groups (non-detectable, 1.5 ± 1.0; Low, 1.6 ± 1.0; Mid, 1.9 ± 1.2; High, 2.1 ± 1.2; P < 0.001), and the proportion of severely frail patients rose from 16.7% in the non-detectable group to 34.2% in the High tertile (P = 0.001 across categories).

### Clinical Outcomes by cFAS and mFI-5

Over the study period, 131 patients (20.0%) died, and 226 (34.5%) experienced a major adverse event. At 1 year, the rates of all-cause mortality were 5.2%, 1.7%, 2.6%, and 9.5% among the non-detectable, Low, Mid, and High cFAS groups, respectively (P = 0.026). Detectable cFAS was associated with significantly higher rates of MAE (38.7% vs. 29.4%; P = 0.011), MALE (13.1% vs. 3.9%; P < 0.001), reintervention (15.9% vs. 10.1%; P = 0.027), and hospital readmission (16.8% vs. 10.1%; P = 0.013). The strongest association was observed for MALE, which occurred in 3.9% of patients with non-detectable cFAS vs. 10.3%, 15.4%, and 13.8% in the Low, Mid, and High tertiles, respectively (P < 0.001). MACE rates did not differ significantly by cFAS level (P = 0.621) (**Table 2**).

**Table 2.** Clinical Outcomes Stratified by cFAS Detectability and cFAS Tertile.

| Outcome | Total<br>(N=657) | Non-Detectable<br>cFAS<br>(n = 306) | Detectable cFAS, pg/mg (n = 351) |  |  | P<br>(4-grp) | P<br>(det) |
| --- | --- | --- | --- | --- | --- | --- | --- |
|  |  |  | Low<br>(≤265, n = 117) | Mid<br>(266–748, n = 117) | High<br>(>748, n = 117) |  |  |
| All-Cause Mortality |  |  |  |  |  |  |  |
| 1-year | 32 (4.9) | 16 (5.2) | 2 (1.7) | 3 (2.6) | 11 (9.5) | 0.026 | 0.697 |
| 3-year | 65 (9.9) | 29 (9.5) | 9 (7.7) | 10 (8.5) | 17 (14.7) | 0.275 | 0.729 |
| 5-year | 92 (14.0) | 40 (13.1) | 15 (12.8) | 14 (12.0) | 23 (19.8) | 0.259 | 0.511 |
| 8-year | 125 (19.1) | 57 (18.6) | 23 (19.7) | 16 (13.7) | 29 (25.0) | 0.178 | 0.794 |
| Overall | 131 (20.0) | 57 (18.6) | 27 (23.1) | 18 (15.4) | 29 (25.0) | 0.219 | 0.421 |
| Composite Morbidity |  |  |  |  |  |  |  |
| MAE | 226 (34.5) | 90 (29.4) | 40 (34.2) | 46 (39.3) | 50 (43.1) | 0.037 | 0.011 |
| MACE | 188 (28.7) | 83 (27.1) | 33 (28.2) | 33 (28.2) | 39 (33.6) | 0.621 | 0.416 |
| MALE | 58 (8.8) | 12 (3.9) | 12 (10.3) | 18 (15.4) | 16 (13.8) | <0.001 | <0.001 |
| Individual Morbidity |  |  |  |  |  |  |  |
| Amputation | 8 (1.2) | 3 (1.0) | 4 (3.4) | 0 (0.0) | 1 (0.9) | 0.121 | 0.730 |
| Reintervention | 87 (13.3) | 31 (10.1) | 15 (12.8) | 21 (17.9) | 20 (17.2) | 0.091 | 0.027 |
| SSI | 24 (3.7) | 10 (3.3) | 5 (4.3) | 5 (4.3) | 4 (3.4) | 0.886 | 0.681 |
| Utilization |  |  |  |  |  |  |  |
| Readmission | 90 (13.7) | 31 (10.1) | 17 (14.5) | 21 (17.9) | 21 (18.1) | 0.071 | 0.013 |

Data are presented as n (%). Outcome data were available for 656 of 657 patients; one patient in the high cFAS tertile with missing follow-up was excluded from event-rate calculations. P (4-grp), comparison across all four groups; P (det), non-detectable vs detectable. MAE, major adverse event; MACE, major adverse cardiovascular event; MALE, major adverse limb event; SSI, surgical site infection; cFAS, circulating fatty acid synthase.

On Kaplan-Meier analysis, cumulative incidence of MAE and MALE increased progressively across cFAS groups, with significant separation of the curves (**Figure 1**; log-rank P = 0.001 and P < 0.001, respectively), whereas curves for MACE and all-cause mortality did not differ significantly by cFAS level. When stratified by mFI-5 category, increasing frailty was associated with progressively worse outcomes for all-cause mortality, MACE, and MAE (**Figure 2**; log-rank P = 0.001, P < 0.001, and P < 0.001, respectively). Cumulative incidence of reintervention rose with higher cFAS (**Supplemental Figure 1**; log-rank P = 0.011) but did not differ significantly across frailty categories. When patients were classified by combined cFAS detectability and frailty status (mFI-5 ≥2), event-free survival separated significantly for MACE, MAE, MALE, and reintervention (**Figure 3**; log-rank P = 0.007, P = 0.001, P < 0.001, and P = 0.005, respectively), with a similar but non-significant trend for all-cause mortality (P = 0.077).

**Figure 1.**
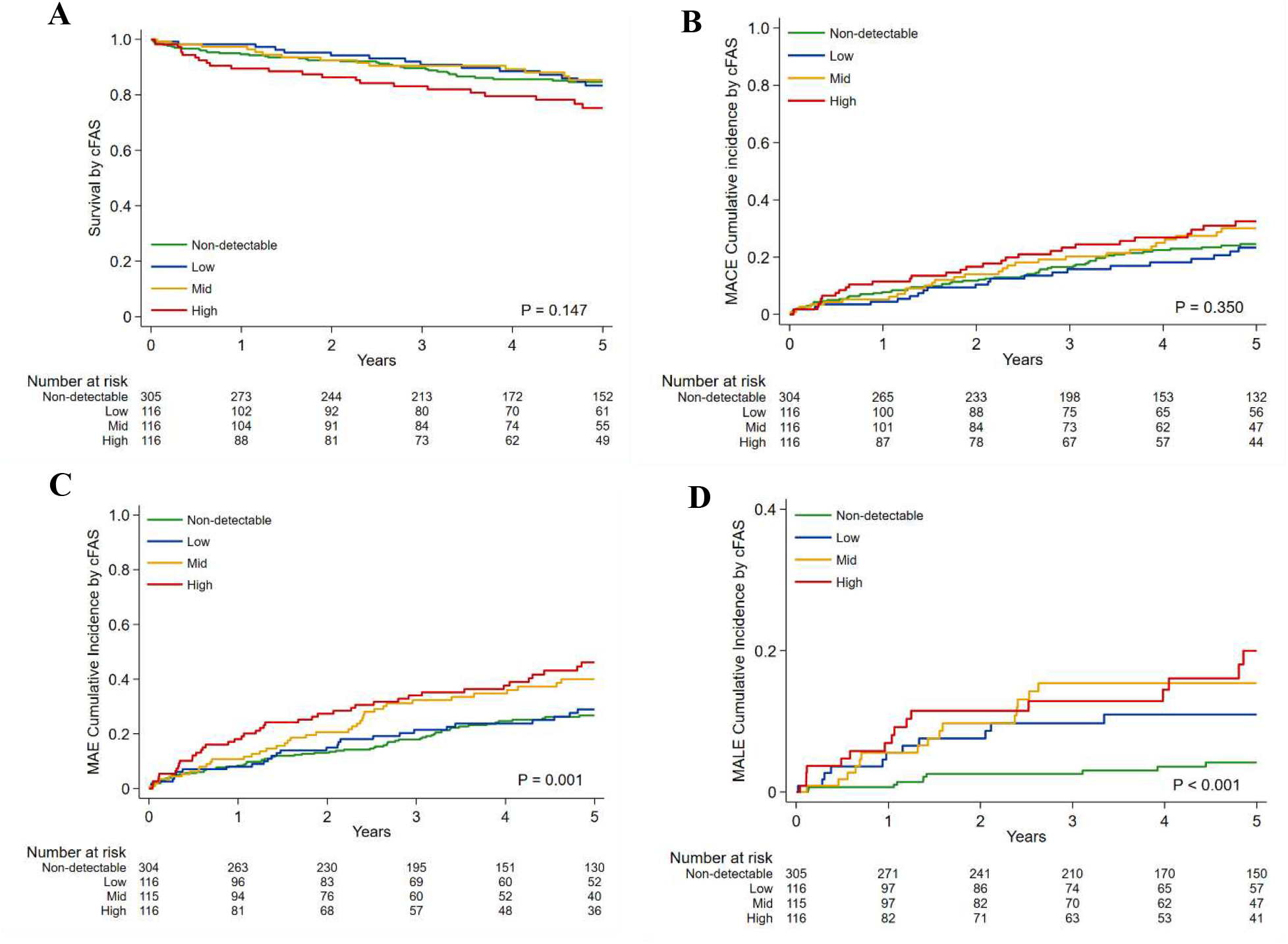
Kaplan-Meier Survival and Cumulative Incidence Curves Stratified by cFAS Level Kaplan-Meier curves for (A) all-cause mortality, (B) major adverse cardiovascular events (MACE), (C) major adverse events (MAE), and (D) major adverse limb events (MALE) stratified by circulating fatty acid synthase (cFAS) level: non-detectable, low (≤265 pg/mg), mid (266–748 pg/mg), and high (>748 pg/mg).

**Figure 2.**
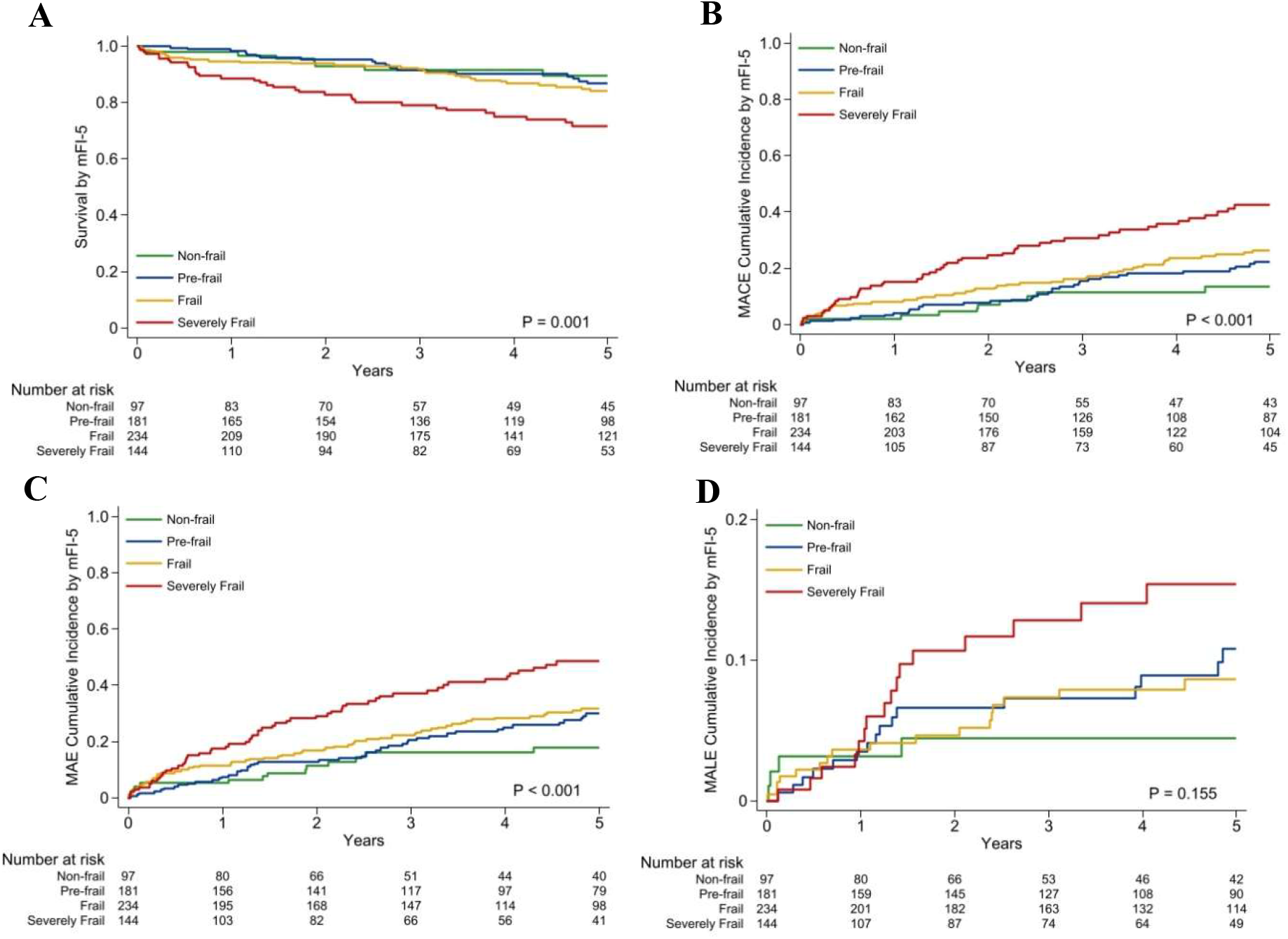
Kaplan-Meier Survival and Cumulative Incidence Curves Stratified by mFI-5 Kaplan-Meier curves for (A) all-cause mortality, (B) major adverse cardiovascular events (MACE), (C) major adverse events (MAE), and (D) major adverse limb events (MALE) stratified by modified frailty index-5 (mFI-5) category: non-frail (score 0), pre-frail (1), frail (2), and severely frail (≥3).

**Figure 3.**
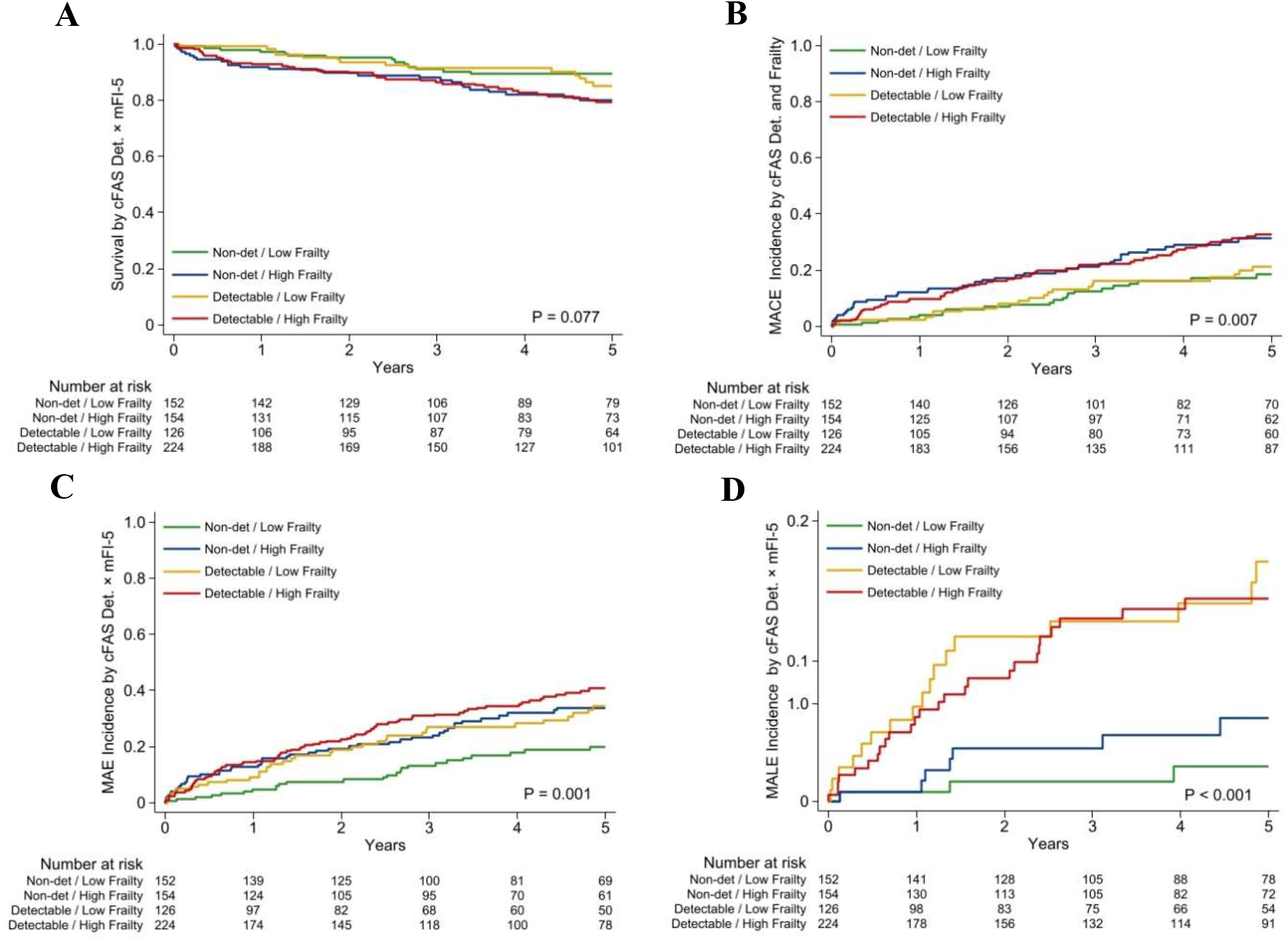
Kaplan-Meier Survival and Cumulative Incidence Curves Stratified by Combined cFAS Detectability and Frailty Status Kaplan-Meier curves for (A) all-cause mortality, (B) major adverse cardiovascular events (MACE), (C) major adverse events (MAE), and (D) major adverse limb events (MALE) stratified by the combination of circulating fatty acid synthase (cFAS) detectability and frailty status (modified frailty index-5 [mFI-5] ≥2): non-detectable cFAS with low frailty, non-detectable cFAS with high frailty, detectable cFAS with low frailty, and detectable cFAS with high frailty.

### Independent Predictors of Adverse Outcomes

All hazard ratios for cFAS are relative to non-detectable cFAS, and those for mFI-5 are relative to non-frail status (score 0). After adjustment for age, sex, smoking status, eGFR, and BMI, cFAS was most strongly associated with MALE. The hazard was elevated across all detectable tertiles and rose with cFAS level, reaching an adjusted HR (aHR) of 9.39 at 1 year (95% CI 1.94-45.55) and 4.53 at 5 years (2.04-10.05; P < 0.001) in the High tertile. cFAS was also independently associated with reintervention, with an aHR in the High tertile of 3.14 at 1 year (1.28-7.68) and 2.50 at 5 years (1.37-4.57) (**Table 3**).

**Table 3.** Cox Proportional Hazards Analysis for Clinical Outcomes by cFAS and mFI-5.

|  | Short-term (1-Year) |  | Long-term (5-Year) |  |
| --- | --- | --- | --- | --- |
|  | HR (95% CI) | aHR (95% CI) | HR (95% CI) | aHR (95% CI) |
| **Major Adverse Events** |  |  |  |  |
| **cFAS, pg/mg** |  |  |  |  |
| Low ($\leq 265$) | 0.95 (0.45–2.05) | 1.01 (0.47–2.19) | 1.09 (0.70–1.69) | 1.13 (0.72–1.76) |
| Mid (266–748) | 1.28 (0.65–2.56) | 1.36 (0.68–2.75) | **1.63 (1.10–2.41)\*** | **1.67 (1.12–2.49)\*** |
| High ($> 748$) | **2.21 (1.22–4.02)\*\*** | **2.20 (1.20–4.06)\*** | **1.98 (1.35–2.90)\*\*\*** | **1.94 (1.31–2.85)\*\*** |
| **mFI-5** |  |  |  |  |
| Pre-frail (1) | 1.35 (0.48–3.80) | 1.02 (0.34–3.05) | 1.59 (0.87–2.89) | 1.27 (0.68–2.37) |
| Frail (2) | 2.05 (0.78–5.37) | 1.59 (0.56–4.55) | 1.76 (0.98–3.14) | 1.38 (0.74–2.55) |
| Severely frail ($\geq 3$) | **3.37 (1.28–8.87)\*** | 2.60 (0.89–7.57) | **3.14 (1.75–5.64)\*\*\*** | **2.46 (1.30–4.65)\*\*** |
| **Major Adverse Cardiovascular Events** |  |  |  |  |
| **cFAS, pg/mg** |  |  |  |  |
| Low ($\leq 265$) | 0.57 (0.21–1.49) | 0.60 (0.23–1.59) | 0.90 (0.55–1.46) | 0.92 (0.56–1.50) |
| Mid (266–748) | 0.68 (0.28–1.68) | 0.74 (0.30–1.85) | 1.22 (0.78–1.89) | 1.25 (0.80–1.96) |
| High ($> 748$) | 1.47 (0.73–2.95) | 1.50 (0.73–3.06) | 1.37 (0.89–2.12) | 1.37 (0.88–2.13) |
| **mFI-5** |  |  |  |  |
| Pre-frail (1) | 1.85 (0.38–8.90) | 1.27 (0.24–6.68) | 1.63 (0.80–3.30) | 1.22 (0.59–2.52) |
| Frail (2) | 3.65 (0.84–15.80) | 2.77 (0.58–13.10) | **2.01 (1.02–3.97)\*** | 1.48 (0.72–3.01) |
| Severely frail ($\geq 3$) | **7.44 (1.74–31.85)\*\*** | **5.58 (1.17–26.73)\*** | **3.78 (1.91–7.46)\*\*\*** | **2.69 (1.29–5.58)\*\*** |
| **All-Cause Mortality** |  |  |  |  |
| **cFAS, pg/mg** |  |  |  |  |
| Low ($\leq 265$) | 0.33 (0.07–1.43) | 0.37 (0.08–1.64) | 0.97 (0.54–1.76) | 1.02 (0.56–1.86) |
| Mid (266–748) | 0.49 (0.14–1.68) | 0.60 (0.17–2.11) | 0.90 (0.49–1.65) | 0.96 (0.52–1.79) |
| High ($> 748$) | 1.96 (0.91–4.22) | 2.21 (0.99–4.92) | **1.68 (1.00–2.80)\*** | **1.77 (1.05–3.00)\*** |
| **mFI-5** |  |  |  |  |
| Pre-frail (1) | 0.79 (0.13–4.72) | 0.48 (0.06–3.78) | 1.17 (0.51–2.65) | 0.85 (0.36–1.99) |
| Frail (2) | 2.53 (0.57–11.29) | 2.14 (0.40–11.45) | 1.45 (0.67–3.17) | 1.04 (0.46–2.38) |
| Severely frail ($\geq 3$) | **5.50 (1.26–24.04)\*** | 4.66 (0.86–25.30) | **3.00 (1.38–6.49)\*\*** | 2.13 (0.92–4.91) |
| **Major Adverse Limb Events** |  |  |  |  |
| **cFAS, pg/mg** |  |  |  |  |
| Low ($\leq 265$) | **8.04 (1.62–39.86)\*** | **8.24 (1.65–41.14)\*** | **3.01 (1.28–7.09)\*** | **3.02 (1.27–7.14)\*** |
| Mid (266–748) | **8.01 (1.62–39.70)\*** | **7.57 (1.52–37.78)\*** | **4.19 (1.88–9.32)\*\*\*** | **3.95 (1.77–8.84)\*\*** |
| High ($> 748$) | **10.46 (2.17–50.37)\*\*** | **9.39 (1.94–45.55)\*\*** | **5.15 (2.34–11.35)\*\*\*** | **4.53 (2.04–10.05)\*\*\*** |
| **mFI-5** |  |  |  |  |
| Pre-frail (1) | 1.03 (0.26–4.13) | 0.97 (0.22–4.34) | 1.93 (0.64–5.77) | 2.03 (0.64–6.46) |
| Frail (2) | 0.97 (0.25–3.75) | 0.92 (0.20–4.12) | 1.54 (0.51–4.61) | 1.67 (0.51–5.44) |
| Severely frail ($\geq 3$) | 1.21 (0.29–5.07) | 1.17 (0.23–5.82) | 2.95 (0.99–8.83) | **3.52 (1.06–11.71)\*** |
| **Reintervention** |  |  |  |  |
| **cFAS, pg/mg** |  |  |  |  |
| Low ($\leq 265$) | 2.15 (0.85–5.44) | 2.13 (0.80–5.69) | 1.40 (0.72–2.74) | 1.40 (0.69–2.81) |
| Mid (266–748) | 2.14 (0.85–5.43) | 2.32 (0.91–5.93) | **1.87 (1.01–3.46)\*** | **1.93 (1.03–3.62)\*** |
| High ($> 748$) | **3.01 (1.25–7.22)\*** | **3.14 (1.28–7.68)\*** | **2.56 (1.42–4.60)\*\*** | **2.50 (1.37–4.57)\*\*** |
| **mFI-5** |  |  |  |  |
| Pre-frail (1) | 1.03 (0.39–2.75) | 1.47 (0.47–4.64) | 1.77 (0.81–3.88) | 2.09 (0.86–5.06) |
| Frail (2) | 0.69 (0.25–1.90) | 1.07 (0.31–3.70) | 0.97 (0.43–2.20) | 1.25 (0.49–3.18) |
| Severely frail ($\geq 3$) | 0.96 (0.33–2.77) | 1.55 (0.42–5.70) | 1.64 (0.71–3.77) | 2.31 (0.88–6.08) |

1 Reference categories: cFAS non-detectable; mFI-5 non-frail (score 0). Covariates: age, sex, smoking status, eGFR, and BMI. HR, 2 hazard ratio; aHR, adjusted hazard ratio; CI, confidence interval; MAE, major adverse events; MACE, major adverse 3 cardiovascular events; MALE, major adverse limb events; mFI-5, modified frailty index-5; cFAS, circulating fatty acid synthase. \*P < .05; \*\*P < .01; \*\*\*P < .001.

For the composite MAE endpoint, High cFAS was associated with a higher hazard at 1 year (aHR 2.20; 95% CI 1.20-4.06) and 5 years (aHR 1.94; 95% CI 1.31-2.85). High cFAS was also associated with 5-year all-cause mortality (aHR 1.77; 95% CI 1.05-3.00), with a similar trend at 1 year (aHR 2.21; 95% CI 0.99-4.92; P = 0.055) (**Table 3**).

mFI-5 showed the opposite pattern and was the stronger predictor of cardiovascular outcomes. Severely frail patients (mFI-5 ≥ 3) had higher 5-year hazards of MAE (aHR 2.46; 95% CI 1.30–4.65) and MACE (aHR 2.69; 95% CI 1.29–5.58) but no association with reintervention; among limb outcomes, only severely frail patients had elevated 5-year MALE risk (aHR 3.52; 95% CI 1.06–11.71) (**Table 3**).

### Added Value for Risk Discrimination

The addition of cFAS and mFI-5 to standard clinical covariates improved discriminatory performance across all endpoints (**Table 4**). The largest gains occurred for limb-specific outcomes. For MALE, cFAS alone increased the C-statistic from 0.649 to 0.764 (ΔC = +0.116) at 1 year and from 0.616 to 0.705 (ΔC = +0.089) at 5 years. The combined model further improved discrimination to 0.768 (ΔC = +0.119) at 1 year and 0.715 (ΔC = +0.098) at 5 years. For reintervention, cFAS alone yielded a ΔC of +0.074 at 1 year, and the combined model reached +0.078.

**Table 4.**
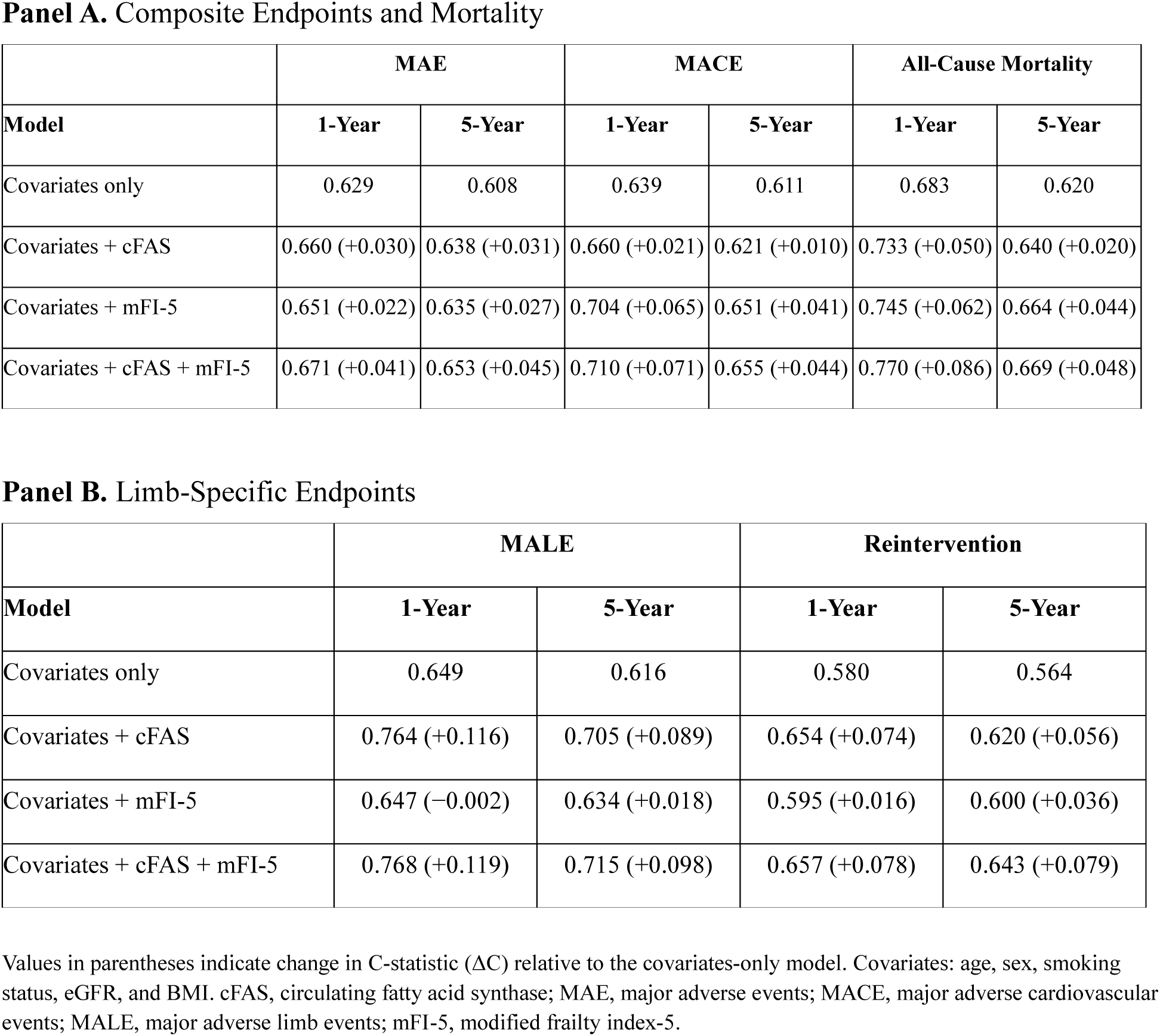
Incremental Discriminatory Value of cFAS and mFI-5 (Harrell’s C-Statistic)

**Panel A.** Composite Endpoints and Mortality
| Model | MAE |  | MACE |  | All-Cause Mortality |  |
| --- | --- | --- | --- | --- | --- | --- |
|  | 1-Year | 5-Year | 1-Year | 5-Year | 1-Year | 5-Year |
| Covariates only | 0.629 | 0.608 | 0.639 | 0.611 | 0.683 | 0.620 |
| Covariates + cFAS | 0.660 (+0.030) | 0.638 (+0.031) | 0.660 (+0.021) | 0.621 (+0.010) | 0.733 (+0.050) | 0.640 (+0.020) |
| Covariates + mFI-5 | 0.651 (+0.022) | 0.635 (+0.027) | 0.704 (+0.065) | 0.651 (+0.041) | 0.745 (+0.062) | 0.664 (+0.044) |
| Covariates + cFAS + mFI-5 | 0.671 (+0.041) | 0.653 (+0.045) | 0.710 (+0.071) | 0.655 (+0.044) | 0.770 (+0.086) | 0.669 (+0.048) |

**Panel B.** Limb-Specific Endpoints
| Model | MALE |  | Reintervention |  |
| --- | --- | --- | --- | --- |
|  | 1-Year | 5-Year | 1-Year | 5-Year |
| Covariates only | 0.649 | 0.616 | 0.580 | 0.564 |
| Covariates + cFAS | 0.764 (+0.116) | 0.705 (+0.089) | 0.654 (+0.074) | 0.620 (+0.056) |
| Covariates + mFI-5 | 0.647 (−0.002) | 0.634 (+0.018) | 0.595 (+0.016) | 0.600 (+0.036) |
| Covariates + cFAS + mFI-5 | 0.768 (+0.119) | 0.715 (+0.098) | 0.657 (+0.078) | 0.643 (+0.079) |

Values in parentheses indicate change in C-statistic ( $\Delta C$ ) relative to the covariates-only model. Covariates: age, sex, smoking status, eGFR, and BMI. cFAS, circulating fatty acid synthase; MAE, major adverse events; MACE, major adverse cardiovascular events; MALE, major adverse limb events; mFI-5, modified frailty index-5.

For cardiovascular endpoints, mFI-5 contributed more than cFAS. For MACE, mFI-5 alone improved the C-statistic by +0.065 at 1 year, compared with +0.021 for cFAS alone. For all-cause mortality, the combined model achieved the highest discrimination at 1 year (C = 0.770; ΔC = +0.086).

### Predictor Independence and Robustness

cFAS and mFI-5 were only weakly correlated (Spearman ρ = 0.19; P < 0.001; n = 351), consistent with the two measures capturing distinct biological constructs. Likelihood ratio tests comparing additive and multiplicative Cox models revealed no significant cFAS × mFI-5 interaction for MAE, MALE, or reintervention at either time horizon (all P > 0.05), consistent with independent and additive contributions to risk prediction. Significant interactions were observed for 5-year MACE (LR χ² = 21.97; P < 0.01) and for all-cause mortality at both time horizons (1-year LR χ² = 11.90, P < 0.01; 5-year LR χ² = 21.74, P < 0.01) (**Supplemental Table 1**).

Sensitivity analyses using alternative predictor specifications were consistent with the primary findings (**Supplemental Table 2**). Modeled as a continuous variable, each 100 pg/mg increase in cFAS was associated with a 3% increase in the 5-year hazard of MAE (aHR 1.03; 95% CI 1.01-1.06; P < 0.05) and a 4% increase in all-cause mortality (aHR 1.04; 95% CI 1.01-1.07; P < 0.05). Log-transformed cFAS was significantly associated with 5-year MAE (aHR 1.25; 95% CI 1.06-1.48; P < 0.01), MACE (aHR 1.22; 95% CI 1.01-1.48; P < 0.05), and all-cause mortality (aHR 1.32; 95% CI 1.03-1.70; P < 0.05). Notably, this continuous association with MACE was not apparent in the categorical analysis, suggesting a graded relationship that the four-level specification did not capture. The categorical specification was retained as primary because no validated clinical cutoffs for cFAS exist and a detectable-versus-non-detectable framework is more readily applied in practice. Each 1-point increase in mFI-5 was associated with a 31% higher 5-year hazard of MAE (aHR 1.31; 95% CI 1.13-1.52; P < 0.001) and a 39% higher hazard of MACE (aHR 1.39; 95% CI 1.18-1.64; P < 0.001). The proportional hazards assumption was satisfied for all adjusted models (all Schoenfeld global test P > 0.05).

## DISCUSSION

Preoperative risk assessment in ASCVD has relied on clinical scores and serum lipids that reflect the burden of cardiovascular risk factors but not the activity of the underlying atherosclerotic disease.^3,4^ To our knowledge, this study is the first to show that a single preoperative measurement of cFAS independently predicts adverse postoperative outcomes after elective vascular surgery. In adjusted models, patients in the highest cFAS group had significantly higher hazards of MAE, MALE, reintervention, and 5-year all-cause mortality. MACE was not predicted by cFAS but was independently predicted by mFI-5, which identified severely frail patients at significantly higher cardiovascular risk. A combined model showed the best discriminative performance across all endpoints. These findings suggest that cFAS and the mFI-5 reflect two complementary aspects of postoperative risk. cFAS reflects disease activity, whereas the mFI-5 reflects physiologic reserve.

Prior work established cFAS as a cross-sectional diagnostic marker of the presence and severity of peripheral atherosclerosis, with concentrations that track plaque FAS content independent of LDL, statin use, diabetes, and smoking, and that distinguish PAD and CLTI from controls.^6,8,10^ In each of these studies, however, cFAS was assessed cross-sectionally as a test for the presence or stage of disease; the present findings extend it from a diagnostic to a prognostic biomarker.

The prognostic signal observed here is consistent with growing evidence that cFAS participates directly in atheroprogression rather than serving as a passive bystander. Human serum containing high concentrations of cFAS promotes macrophage foam-cell formation *in vitro*. In addition, pharmacologic inhibition of FAS, or liver-specific deletion of hepatic *Fasn*, lowers circulating cFAS and reduces aortic atherosclerosis in apolipoprotein E-deficient mice without altering total cholesterol.^9^ At the molecular level, FAS catalyzes the *de novo* synthesis of saturated fatty acids that accumulate in plaque. It also promotes the transformation of macrophages and vascular smooth muscle cells into foam cells and organizes the macrophage plasma membrane for inflammatory signaling.^7,23^ Together, these findings indicate that cFAS is not an innocent bystander and contributes directly to the lipid accumulation and inflammation of the progressing plaque. In the present cohort, cFAS predicted adverse outcomes independent of LDL, statin use, and hyperlipidemia prevalence, a pattern consistent with a marker that reflects lesion-level disease activity rather than systemic lipid burden.

The prognostic contribution of cFAS is best understood in comparison with existing preoperative biomarkers. Clinical risk scores and serum lipids, principally LDL, remain the basis of cardiovascular risk assessment and are recommended in current American Heart Association and American College of Cardiology prevention guidelines.^3^ Their limitations are most evident in treated patients. Among more than 31,000 statin-treated patients across three randomized trials, residual inflammatory risk, but not residual LDL, predicted subsequent cardiovascular events and death.^4^ Once lipids are controlled, the remaining risk reflects ongoing arterial inflammation that LDL does not capture.^4^ This residual inflammatory risk is also modifiable. In the CANTOS trial, anti-inflammatory therapy with canakinumab (a monoclonal antibody targeting interleukin-1β) reduced MACE by 15% relative to placebo without lowering LDL, confirming that atherosclerotic disease activity drives events independently of the lipid burden.^5^ C-reactive protein (CRP), the most established marker of this residual risk, reflects systemic rather than lesion-specific inflammation. Each standard-deviation increase in CRP carries a 37% higher risk of coronary heart disease, yet CRP adds little to risk discrimination beyond conventional factors.^24^ Preoperative natriuretic peptides and perioperative high-sensitivity troponin improve cardiac risk stratification around noncardiac surgery, but they reflect myocardial wall stress and ischemic injury rather than the atherosclerotic disease in the arterial wall.^15,25^ cFAS, by contrast, may more directly reflect the activity of that disease.^6,8^ In this cohort, cFAS rose with diabetes, insulin use, and HbA1c but not with LDL or statin use, indicating that it captures a metabolic dimension of atherosclerotic risk that conventional lipid measures do not currently reflect.

Frailty captured a second, independent dimension of perioperative risk. The mFI-5 predicts postoperative mortality and major complications across surgical specialties and, despite requiring only five routinely documented comorbidities, performs as well as or better than both the Charlson Comorbidity Index and the original 11-item frailty index.^11–13^ In vascular surgery specifically, frailty ranks among the strongest predictors of long-term survival and independently predicts adverse outcomes after abdominal aortic aneurysm repair.^14,26^ In the present study, severely frail patients had roughly 2.5-fold higher 5-year hazards of both MACE and MAE. This is consistent with the graded mortality risk reported in larger vascular surgery cohorts, where the 5-year mortality hazard rises from 2.7-fold with mild frailty to 5.9-fold with moderate-to-severe frailty.^14^ Notably, frailty did not predict reintervention and predicted MALE only in severely frail patients at 5 years, the inverse of the pattern seen for cFAS and consistent with the two measures capturing different dimensions of perioperative risk.

Adding a serum biomarker to clinical risk assessment can improve prediction beyond either component alone. The present results extend this principle to vascular surgery. A preoperative frailty index predicted cardiovascular events after major noncardiac surgery even after accounting for the cardiac biomarker NT-proBNP, much as the mFI-5 retained its predictive value alongside cFAS in the present study. Likewise, combining a panel of inflammatory cytokines (IL-6, IL-10, and CXCL9) with a frailty index improved long-term mortality prediction, with the highest risk in patients elevated on both.^27,28^ Adding cFAS to clinical covariates raised the C-statistic for MALE from 0.649 to 0.764 at 1 year in the present cohort, a larger gain than any other predictor in the model. By comparison, established cardiovascular biomarkers add little once they are layered onto conventional clinical risk factors. In a cohort of more than 240,000 adults, adding CRP or fibrinogen to standard risk factors improved the C-statistic by just 0.004 and 0.003, respectively, and even a panel of ten biomarkers raised it by no more than about 0.01. These gains are roughly an order of magnitude smaller than the 0.12 increase that cFAS provided for MALE in the present cohort.^29,30^ In this cohort, the combined model discriminated best for every endpoint, with the mFI-5 weighted toward cardiovascular outcomes and cFAS toward limb outcomes. Both measures are readily obtained in practice: the mFI-5 from documented comorbidities and cFAS from a single preoperative serum sample.

Several limitations should be considered. The study was retrospective and conducted at a single center, although consecutive enrollment in a prospectively maintained biobank over nearly a decade reduces the likelihood of selection bias. The functional component of the mFI-5 was approximated from neurological and cognitive diagnoses, an approach consistent with the Charlson framework but one that may attenuate the measured association with frailty.^17^ cFAS was measured at a single preoperative time point, and whether its concentration changes after revascularization or with medical therapy is unknown. The cohort was predominantly White and drawn from a single institution, and validation in larger, more diverse, prospective cohorts is needed in the future.

In conclusion, preoperative cFAS and the mFI-5 independently predicted adverse outcomes after elective vascular surgery and captured complementary dimensions of risk. cFAS reflected the activity of the atherosclerotic disease and predicted limb events, whereas the mFI-5 reflected physiologic reserve and predicted cardiovascular events. Together, they improved riskdiscrimination beyond standard clinical assessment. To our knowledge, cFAS is the first serum biomarker of atherosclerotic disease activity shown to predict postoperative outcomes in this population, and prospective external validation is needed before it is adopted as an adjunct to established preoperative risk stratification.

## ACKNOWLEDGEMENTS

This study was supported by the vascular biobank at Washington University School of Medicine in St. Louis. The authors thank the biobank team personnel for their assistance with consenting preoperative patients and collection of fasting blood samples for cFAS analysis. We specifically thank Mrs. Laura McDonald, Mrs. Ashley Cosentino, Mrs. Amanda Penrose, Dr. Connor Engel, Mr. Rodrigo Meade, and Dr. Shahab Hafezi.

## AUTHOR CONTRIBUTIONS

Conception and design: MZ, FG, MAZ

Analysis and interpretation: MZ, WA, FG, MAZ

Data collection: MZ, RC, BK, AE, OS, JY, DI, WA

Writing the article: MZ, MAZ

Critical revision of the article: MZ, FG, MAZ

Final approval of the article: MZ, RC, BK, AE, OS, JY, DI, WA, FG, MAZ

Statistical analysis: MZ, FG

Obtained funding: MAZ

Overall responsibility: MAZ

## DATA AVAILABILITY

The data underlying this article are available in the article and its online supplementary material. Raw data will be shared on reasonable request to the corresponding author.

## Supplemental Materials

**Supplemental Table 1.**
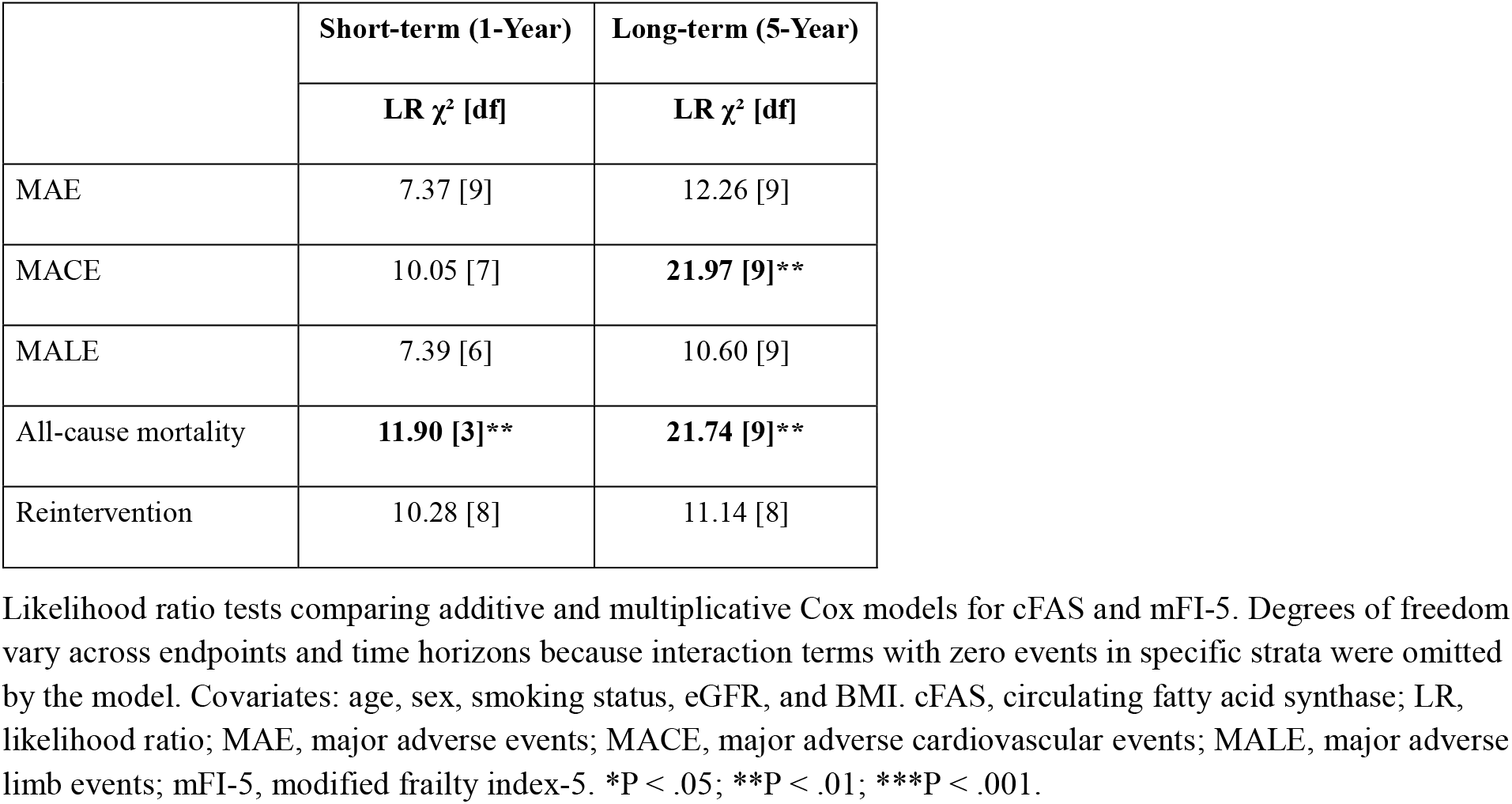
Likelihood Ratio Tests for cFAS × mFI-5 Interaction.

|  | Short-term (1-Year) | Long-term (5-Year) |
| --- | --- | --- |
| | LR $\chi^2$ [df] | LR $\chi^2$ [df] |
| MAE | 7.37 [9] | 12.26 [9] |
| MACE | 10.05 [7] | **21.97 [9]\*\*** |
| MALE | 7.39 [6] | 10.60 [9] |
| All-cause mortality | **11.90 [3]\*\*** | **21.74 [9]\*\*** |
| Reintervention | 10.28 [8] | 11.14 [8] |

3 Likelihood ratio tests comparing additive and multiplicative Cox models for cFAS and mFI-5. Degrees of freedom 4 vary across endpoints and time horizons because interaction terms with zero events in specific strata were omitted 5 by the model. Covariates: age, sex, smoking status, eGFR, and BMI. cFAS, circulating fatty acid synthase; LR, 6 likelihood ratio; MAE, major adverse events; MACE, major adverse cardiovascular events; MALE, major adverse 7 limb events; mFI-5, modified frailty index-5. \*P < .05; \*\*P < .01; \*\*\*P < .001.

**Supplemental Table 2.** Cox Proportional Hazards Analysis with Alternative Predictor Specifications.

|  | Short-term (1-Year) |  | Long-term (5-Year) |  |
| --- | --- | --- | --- | --- |
|  | HR (95% CI) | aHR (95% CI) | HR (95% CI) | aHR (95% CI) |
| **Major Adverse Events** |  |  |  |  |
| cFAS per 100 pg/mg | **1.04 (1.00–1.08)\*** | 1.03 (1.00–1.07) | **1.04 (1.01–1.06)\*\*** | **1.03 (1.01–1.06)\*** |
| ln(cFAS) | 1.31 (0.98–1.75) | 1.28 (0.96–1.71) | **1.29 (1.09–1.53)\*\*** | **1.25 (1.06–1.48)\*\*** |
| mFI-5 score (per point) | **1.45 (1.17–1.80)\*\*** | **1.45 (1.13–1.85)\*\*** | **1.35 (1.18–1.54)\*\*\*** | **1.31 (1.13–1.52)\*\*\*** |
| **Major Adverse Cardiovascular Events** |  |  |  |  |
| cFAS per 100 pg/mg | 1.03 (0.99–1.09) | 1.03 (0.98–1.08) | **1.03 (1.00–1.06)\*** | 1.02 (1.00–1.05) |
| ln(cFAS) | 1.36 (0.92–1.99) | 1.34 (0.91–1.97) | **1.25 (1.03–1.52)\*** | **1.22 (1.01–1.48)\*** |
| mFI-5 score (per point) | **1.76 (1.37–2.27)\*\*\*** | **1.79 (1.34–2.40)\*\*\*** | **1.45 (1.25–1.68)\*\*\*** | **1.39 (1.18–1.64)\*\*\*** |
| **All-Cause Mortality** |  |  |  |  |
| cFAS per 100 pg/mg | **1.06 (1.00–1.11)\*** | **1.06 (1.00–1.11)\*** | **1.04 (1.01–1.07)\*** | **1.04 (1.01–1.07)\*** |
| ln(cFAS) | **1.93 (1.13–3.30)\*** | **1.91 (1.11–3.28)\*** | **1.32 (1.03–1.71)\*** | **1.32 (1.03–1.70)\*** |
| mFI-5 score (per point) | **1.85 (1.36–2.51)\*\*\*** | **1.94 (1.36–2.76)\*\*\*** | **1.46 (1.22–1.76)\*\*\*** | **1.41 (1.15–1.73)\*\*** |
| **Major Adverse Limb Events** |  |  |  |  |
| cFAS per 100 pg/mg | 1.03 (0.98–1.09) | 1.02 (0.97–1.08) | 1.03 (0.98–1.07) | 1.02 (0.98–1.06) |
| ln(cFAS) | 1.12 (0.76–1.65) | 1.05 (0.70–1.57) | 1.18 (0.90–1.53) | 1.13 (0.86–1.48) |
| mFI-5 score (per point) | 1.03 (0.70–1.54) | 1.05 (0.67–1.63) | 1.21 (0.94–1.55) | 1.29 (0.97–1.70) |
| **Reintervention** |  |  |  |  |
| cFAS per 100 pg/mg | 1.03 (0.99–1.08) | 1.03 (0.99–1.08) | 1.03 (0.99–1.07) | 1.02 (0.99–1.06) |
| ln(cFAS) | 1.11 (0.79–1.54) | 1.14 (0.81–1.60) | 1.11 (0.87–1.41) | 1.11 (0.87–1.42) |
| mFI-5 score (per point) | 0.94 (0.69–1.28) | 1.07 (0.75–1.51) | 1.01 (0.81–1.25) | 1.09 (0.86–1.39) |

3 cFAS analyses restricted to patients with detectable cFAS levels (n = 351); mFI-5 analyses performed on all patients with 4 available mFI-5 data. cFAS per 100 pg/mg represents a per-100 unit increase in circulating FAS; ln(cFAS), natural log- 5 transformed cFAS (per 1-unit increase in log scale); mFI-5 score, continuous (per 1-point increase). Covariates: age, sex, 6 smoking status, eGFR, and BMI. HR, hazard ratio; aHR, adjusted hazard ratio; CI, confidence interval; MAE, major adverse 7 events; MACE, major adverse cardiovascular events; MALE, major adverse limb events; mFI-5, modified frailty index-5; cFAS, 8 circulating fatty acid synthase. \*P < .05; \*\*P < .01; \*\*\*P < .001.

**Supplemental Figure 1.**
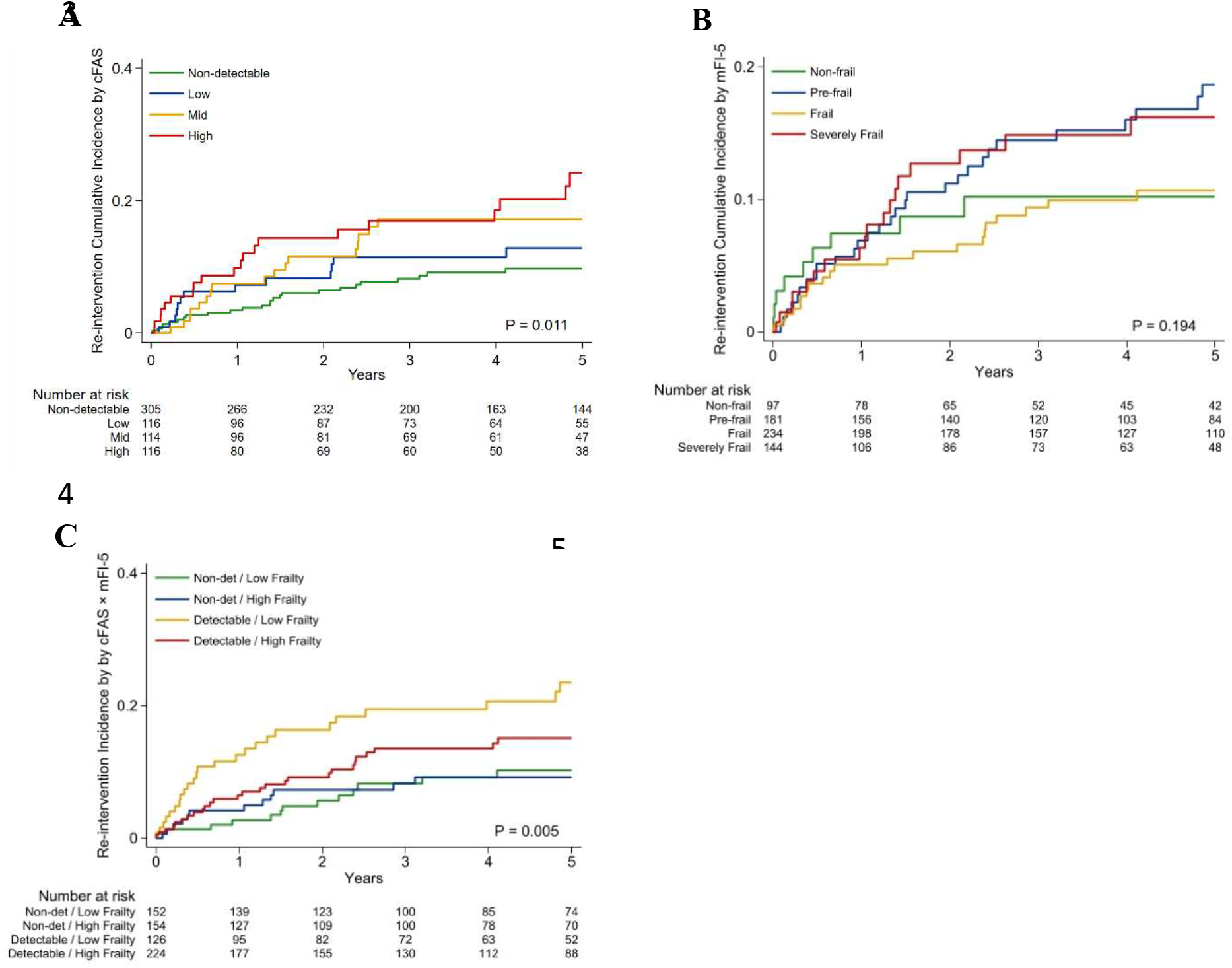
Kaplan-Meier Cumulative Incidence of Reintervention Stratified by cFAS, mFI-5, and Combined cFAS Detectability–Frailty Status Cumulative incidence of target-related reintervention stratified by (A) circulating fatty acid synthase (cFAS) level: non-detectable, low (≤265 pg/mg), mid (266–748 pg/mg), and high (>748 pg/mg); (B) modified frailty index-5 (mFI-5) category: non-frail (score 0), pre-frail (1), frail (2), and severely frail (≥3); and (C) combined cFAS detectability and frailty status: non-detectable cFAS with low frailty (mFI-5 <2), non-detectable cFAS with high frailty (mFI-5 ≥2), detectable cFAS with low frailty, and detectable cFAS with high frailty.

